# Quantifying User Engagement with the Helpilepsy Electronic Seizure Diary

**DOI:** 10.64898/2026.08.05.26359796

**Authors:** Joe Davies, Andrea Biondi, Pedro Viana, Ludovic Ampe, Jonathan Schreiber, Mark P Richardson

## Abstract

Seizure diaries are one of the most useful sources of information in the management of epilepsy, however patient engagement with them can be sporadic. Sustained participation with seizure diaries affects the completeness and reliability of self-reported data, so it is vital to be able to measure engagement. To facilitate this, we create a multidimensional engagement metric with which to characterize how patients interact with their seizure diary. We utilise data from the Helpilepsy, a seizure diary application, common features found in application engagement metrics in business settings, and well understood clinical features to do this. Clustering is then performed to isolate different user groups based on how engaged they are, and these groups are studied to understand what drives the differences in engagement.

We found three groups emerge from the clustering: low, medium and highly engaged users. Investigating these groups further, we put together a “profile” for highly-engaged users. We find that they tend to be older at the point of diagnosis, and have had epilepsy for longer than the other users. We also find they tend to have had more medications, have higher doses of common anti-seizure medications, and they have more medications typically given to those with refractory epilepsy.

The implications for e-diary design are that more attention should be given to those newer to epilepsy in the onboarding phase. Also, engagement is not necessarily based on just the upload of seizures, with other features of an e-diary being important to be filled in.

## I. Introduction

APPROXIMATELY one-third of people with epilepsy continue to experience seizures despite appropriate antiseizure medication treatment, making accurate seizure monitoring a fundamental component of long-term management [1]. Reliable seizure documentation is essential for evaluating treatment response, identifying seizure triggers and clustering, and guiding therapeutic decisions, while also helping patients and caregivers manage the uncertainty associated with the unpredictable nature of seizures [2].

Patients recognize the value of seizure documentation. In a survey by Blachut et al [3], 77% of people with epilepsy considered seizure recording important for monitoring seizure frequency, following physician recommendations, and identifying potential triggers. However, participants may over-report their seizure or their retrospective recall can be incomplete, particularly for seizures occurring during sleep or associated with impaired awareness [4] [5] [6] [7]. These limitations have driven increasing interest in digital tools designed to facilitate more structured and consistent seizure tracking and to support sustained patient engagement [8].

Over the past decade, paper diaries have largely been replaced by electronic seizure diaries and smartphone applications that enable users to record seizures, medications, sleep, mood, and potential triggers while sharing data with clinicians and care-givers. A systematic scoping review identified more than 10 such platforms and reported generally high patient interest and acceptable adherence, particularly when integrated into clinical care [8]. Beyond seizure counting, self-reported data from platforms such as My Seizure Diary (www.myseizurediary.com), Seer Medical (www.seermedical.com), and SeizureTracker (www.seizuretracker.com) have been also widely used to study seizure clustering, temporal patterns, and forecasting, but rarely to characterize multidimensional engagement.

Diary data have been used to identify seizure clusters [9], characterize circadian and multiday seizure cycles [10] [11], investigate associations between seizures and triggers such as stress, sleep, and mood [12], and develop personalized seizure forecasting models [11]. More broadly, self-reported variables are increasingly recognized as a valuable source of information for epilepsy monitoring and management, particularly when integrated with wearable or implantable technologies [8].

Several studies have also demonstrated that electronic seizure diaries can be used consistently over periods ranging from several months to more than one year, although adherence and retention vary substantially across individuals and study designs. In structured research settings, adherence has ranged from approximately 55% to more than 80%, depending on the frequency of required entries, reminder systems, and integration with clinical care [12] [13] [14] [15] [16]. In a large observational cohort of 560 people with epilepsy using Helpilepsy®, retention was 71% at 3 months and 58% at 6 months when retention was defined as at least one weekly use. Importantly, users who shared data with healthcare professionals demonstrated substantially higher retention than standalone users, emphasizing the importance of clinical integration in sustaining long-term engagement [17].

Recent studies combining seizure diaries with wearable and electroencephalographic monitoring further support the feasibility of long-term diary use. Similarly, Biondi et al. [15] reported a high level of acceptability for the use of seizure diary over 6 months by 12 patients with epilepsy. Additionally, feasibility studies have shown moderate interest (31.8 to 50%) in continuing app use after study completion [15] [18].

Although all these studies have provided valuable information on feasibility, adherence and retention, these measures capture only a limited aspect of user engagement. Most investigations have been conducted in structured settings, such as clinical trials, feasibility studies, or hospital-based monitoring, and have focused primarily on whether participants continued using an application rather than on how they interacted with its different features over time. As a result, an important knowledge gap remains regarding the multidimensional nature and longitudinal evolution of engagement with electronic seizure diaries in routine clinical practice.

In epilepsy, patient engagement has been identified as an important determinant of performance expectancy and the successful implementation of digital health interventions [19]. A more comprehensive understanding of engagement is particularly relevant given the marked heterogeneity of the disorder. Patients differ substantially in seizure frequency, seizure awareness, treatment complexity, disease duration, and comorbidities, and their motivations for using a seizure diary are likely to vary accordingly. Characterizing patterns of engage-ment may therefore reveal clinically meaningful differences in patient needs, motivations, and self-management behaviours. Understanding engagement is also critical because sustained participation directly affects the completeness and reliability of self-reported data. Reduced engagement may result in missing or inconsistent seizure, sleep, and medication records, limiting the usefulness of these data for clinical decision-making, seizure forecasting, and the development of personalized digital health tools.

To address this knowledge gap, we aimed to develop and apply a multidimensional engagement framework to characterize how people with epilepsy interact with the Helpilepsy® electronic seizure diary. The availability of a large, anonymized, longitudinal dataset comprising more than 20,000 unique users and including information on seizure reports, sleep scores, medication reminders, and profile completion provides a unique opportunity to examine engagement at scale in a real-world setting.

Specifically, we aimed to:

1. Construct a composite engagement score incorporating profile completeness ratio, seizure and sleep reporting regularity, longitudinal use of optional features, and medication reminder utilization.
2. Identify distinct engagement groups using unsupervised clustering methods.
3. Evaluate the stability of engagement over time using expanding and fixed temporal windows.
4. Compare demographic and clinical characteristics across engagement groups, including age, age at diagnosis, epilepsy duration, and medication burden.
5. Explore whether highly engaged users represent a clinically distinct subgroup, potentially reflecting individuals with greater treatment complexity and stronger motivation for self-management.

By quantifying engagement in a large real-world epilepsy cohort, this study aims to improve understanding of how patients use electronic seizure diaries and to inform the design and development of future digital health tools and personalized epilepsy monitoring.

## II. Method

### A. Study Information

This study involved secondary analysis of pre-existing deidentified data collected through the Helpilepsy® mobile application (Neuroventis BV, Overijse, Belgium). Users of the application were provided with an explicit opt-in option to allow their health data to be used in anonymized form for scientific research, as described in the Neuroventis privacy policy (Supplementary Material 1). Only data from users who provided this consent were eligible for inclusion. Data from individuals participating in interventional clinical trials or other sponsored research studies were excluded.

Before transfer to King’s College London, Neuroventis irreversibly de-identified the dataset by removing direct identifiers, free-text fields, and other potentially identifying information. The transfer and processing of the data were governed by a Data Sharing Agreement between Neuroventis BV and King’s College London (King’s reference 2844623), signed in July 2024, for non-commercial research purposes and in compliance with UK GDPR and EU GDPR.

Because the study involved analysis of existing de-identified data and did not involve recruitment, intervention, or access to identifiable personal information, additional Research Ethics Committee approval was not required under applicable UK research governance regulations.

### B. Helpilepsy App

Helpilepsy® (Neuroventis BV, Overijse, Belgium) is a CE-marked digital platform designed to support self-management and clinical monitoring in epilepsy. The platform consists of a patient-facing mobile application and a web-based dashboard for healthcare professionals. Through the mobile application, users can record seizure events, antiseizure medications, side effects, sleep, mood, appointments, and potential seizure triggers, and can receive medication reminders and personalized reports summarizing their epilepsy history. Data entered by patients can be securely shared with treating clinicians, who can review longitudinal trends through an interactive dashboard to support clinical decision-making. The platform can also integrate data from compatible seizure detection devices to complement self-reported information (www.helpilepsy.com). Data analyzed in this study were collected through the Helpilepsy application between 2017 and 2026. The dataset comprised de-identified longitudinal records from 25,782 unique users who had explicitly consented to the use of their anonymized data for scientific research. Users participating in clinical trials or other sponsored research studies were excluded. Available variables included demographic and clinical characteristics (e.g., age, sex, country, and age at epilepsy onset), seizure reports and their timestamps, medication information, side effects, medication reminders, and self-reported mood and sleep scores. Mood and sleep were recorded using 11-point Likert scales ranging from 0 to 10 inclusive, with higher values reflecting greater self-reported intensity or severity. No direct personal identifiers were included in the dataset.

### C. Analysis

Data are stored on encrypted, password-protected King’s College London servers and, when temporarily accessed on local devices, are maintained on encrypted, password-protected hard drives. All analyses were conducted using Python (Python Software Foundation, Wilmington, DE, USA).

### D. Engagement Metric

In order to measure the engagement of a user with a seizure diary application, we need to consider how a user that is “engaged” is likely to act. In industry, metrics such as: the daily to monthly active user ratio [20], click-through rate [21], and feature adoption rate [22] consider not just whether a user spends time on an app, but also what they do when they are using it. These focus on actions that users take by choice, as well as those that happen naturally whilst navigating the app.

We focus on the same type of actions in this research. Our metric is defined as:

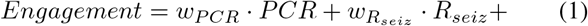

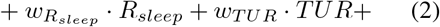

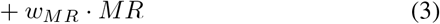

where: *PCR* is profile completeness ratio, *R*_*seiz*_ and *R*_*sleep*_ are seizure and sleep upload regularity respectively, *TUR* is the tenure usage ratio (of sleep scores in this case), and *MR* is whether someone has a medication reminder set. The *w*_*X*_ values are the associated weights that follow:

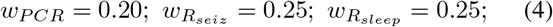

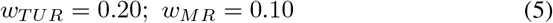

such that the sum of the weights is 1.

Profile completeness ratio (PCR) refers to the percentage of possible features collected by the app (mood score, gender, birth year etc) that a user has filled in. This gives an indication of how many of the features the user has engaged with. This is calculated by:

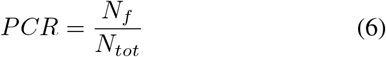

where: *N*_*f*_ is the number of features with at least one input and *N*_*tot*_ is the total number of features.

Seizure and sleep score upload regularity are used to measure not just how many of each a user will upload, but also whether this is kept up regularly over time. Regularity is calculated by first calculating the normalised “burstiness” score [23]. Burstiness, in this work, refers to how clustered uploads of seizures or sleep scores are. We find this by getting the standard deviation (sig) and mean (mu) of the times between seizure uploads and calculating:

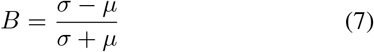

where *B* is burstiness. Those users with 3 or fewer uploads of seizures or sleep have *B* set to 0 to avoid divergences that may lead to unrealistic results. Calculating the mean or variance of fewer than 3 values is not computable. Burstiness is typically between -1 and 1 in value. To normalise to 0 and 1, we calculate:

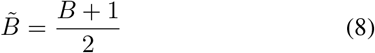

where 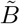 is the normalised burstiness.

Burstiness can be considered a measure of irregularity, so to find regularity we calculate:

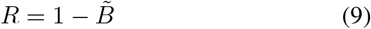

However, there is a conceptual problem with this approach. Whilst we do want to reward those that upload more scores, we do not wish to penalise those people that are regular with their uploads. Users that do not have many seizures, but regularly upload the ones they believe themselves to have had, are still engaged users. To combat this, a saturation function is added to the regularity score, calculated as:

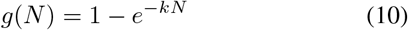

where: N is the number of uploads and k is a saturation constant which dictates how fast the function levels off. A higher k-value confers a steeper saturation curve. For this work, we note that the average number of uploads of both seizures and sleep scores is low, so a relatively high k-value is chosen of 0.4. Regularity is thus fully defined as:

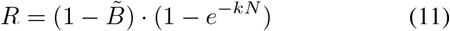

This same definition is used for both sleep score and seizure uploads, with the change coming from the relative standard deviations, means and number of samples in the distributions. Tenure Usage Ratio (TUR) for sleep scores is defined as the ratio of number of unique days scores were uploaded on vs the total number of days a user has had the app. It is defined as:

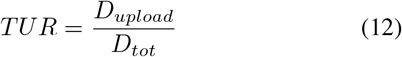

where *D*_*upload*_ is the number of unique days something has been uploaded and *D*_*tot*_ is the total number of days on the app.

TUR is often used in application engagement metrics to identify an user’s tenure with an application, and whether they tend to keep using the app actively over time. For our purposes, we leverage the same idea to identify whether a user is engaged over time with an optional component of the app (sleep score uploads) that does not have a biological trigger to push them to upload something i.e. a seizure being identified.

Finally, medication reminder (MR) is either a 0 or 1 and is simply defined as whether the user has medication reminders present. This is added to act as a proxy for whether a user is active in using the app to manage a given treatment for their epilepsy.

The weights are chosen such that we give equal importance to seizure uploads and sleep score upload regularity. Being a regular uploader is a clear indicator of engagement with the app. MR is weighted lower than the rest, but not as an indicator of importance. This is to mitigate the binary nature of the component and to stop it from dominating the overall engagement score.

The engagement score is calculated per user and outliers are removed using the inter-quartile range (IQR) method. Unphysical results — those falling out of the range [0,1] — are removed. These can arise from those with very low uploads, which force some metric components to blow up to unreasonably high values. Also, those values coming from users that supposedly upload unphysical numbers of seizures (of the order of hundreds per day), are also removed. After these operations are completed, we are left with engagement scores for a total of 17,151 users.

### E. Clustering

The next stage in the analysis is to understand whether we can identify differences between users in the data. To investigate this, we cluster the features making up the engagement metric using an elbow plot [24]. The elbow method works by first defining a range of cluster numbers, k, typically [1, 10]. Then for each k, KMeans Clustering [25] is used. This randomly initialises k centroids in the data and measures the distance to that centroid from each datapoint, updating the centroid to be equidistant from each, defining a cluster. Using KMeans we can then calculate the sum of squared-errors, SSE, i.e. how close the data points are to their centroids overall. We then plot the SSE for each k. This can be found in figure 1. In order to find the number of clusters, a visual inspection of the slope is made that finds where the gradient significantly flattens out. In figure 2 this is clear at 3 clusters, though also could be said to happen at 4. To confirm either of these, we also calculate a silhouette score [26] is calculated for both 3 and 4 clusters. A silhouette score is a measure of how similar each data point is to its own cluster vs other clusters and is found by:

**Fig. 1:**
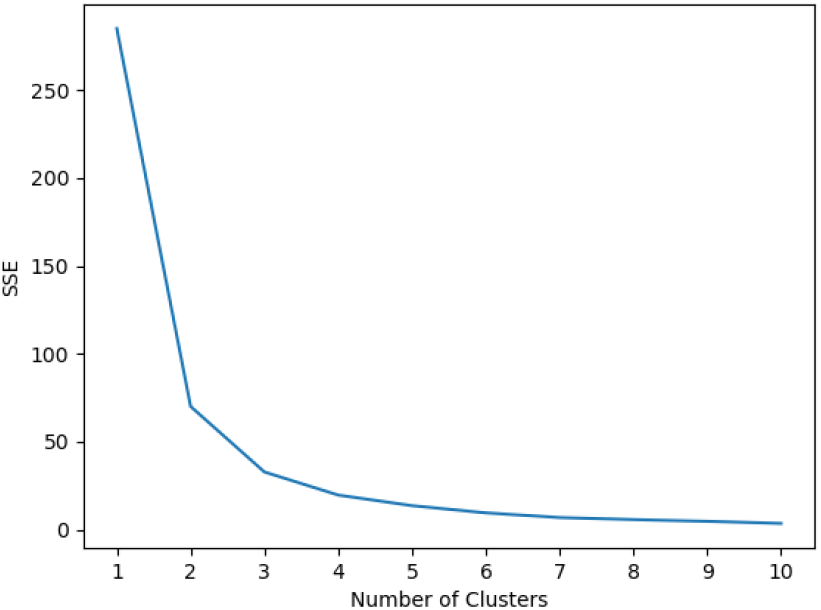
Elbow plot for our data. There is an indication of 3 being the correct number of clusters, though 4 is also viable.

**Fig. 2:**
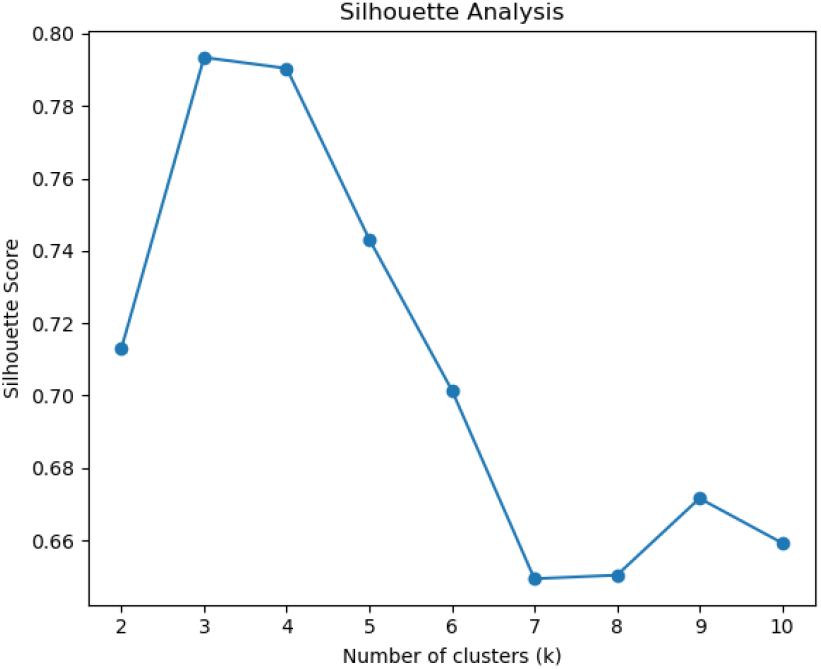
The silhouette scores for each cluster. 3 clusters gives the highest result.

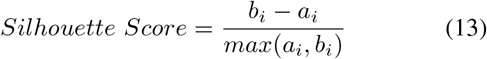

where: *a*_*i*_ is intra-cluster distance, and *b*_*i*_ is inter-cluster distance for each point i. A high score indicates that a point is well classified by a cluster. Getting an overall measure of silhouette score for a set of k clusters shows how well that number of clusters describes the data. A plot of the silhouette scores for each cluster number is found in figure 2. We can see from this that 3 clusters gives the highest value, strengthening the argument for 3 clusters. In addition, the value for 3 clusters being close to 0.8 is high, showing that this number of clusters describes the data well. A histogram of user engagement scores is shown in figure 3. This shows the three clusters clearly. Classifying each user into a cluster number allows us to define boundaries for each cluster. We can name them as low, medium and high engagement groups, defined in table I. We see a clear delineation between low, medium and highly engaged users. It should be noted that the weighting can affect the distribution of engagement scores. To test whether these clusters are still present in the engagement after reasonable reweighting, we test two scenarios. The first is re-weighting such that all components are treated equally i.e. all weights are set to 0.2. This can be seen in figure 4a. The second is when we increase the weights on the components that come from non-biologically triggered events i.e. not from seizures or medication to combat seizures. In this regime we have weightings from TUR, *R*_*sleep*_ and PCR all being 0.25, MR being 0.1 and *R*_*seiz*_ being 0.15. The engagement score distribution for this can be seen in figure 4b. It is clear from both that close to the same distribution is present, showing the groups survive re-weighting and follow approximately the same boundaries.

**Fig. 3:**
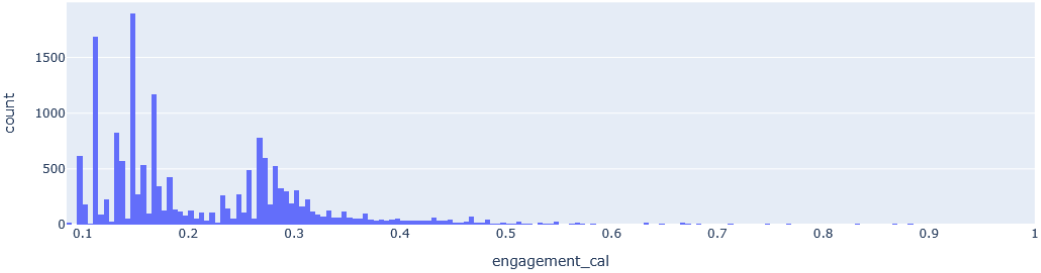
Histogram of engagement over all users. This shows a clear multi-modal distribution, indicative of distinct clusters.

**Fig. 4:**
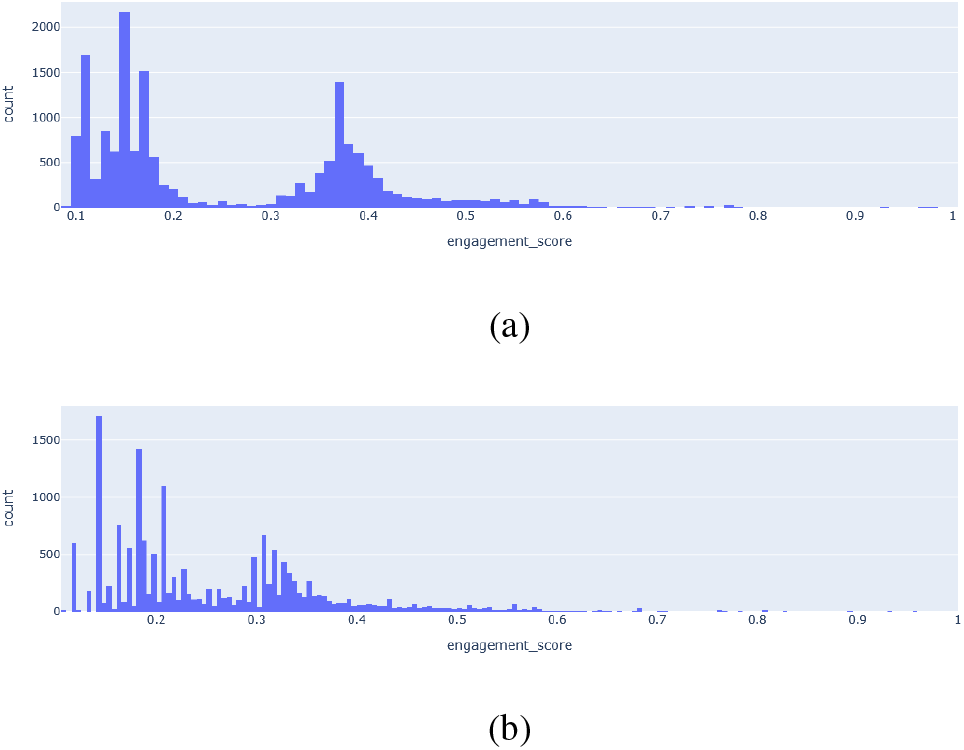
Engagement score distributions for reweightings of the components: (a) equal weights and (b) focusing on optional-type uploads.

**TABLE I:**
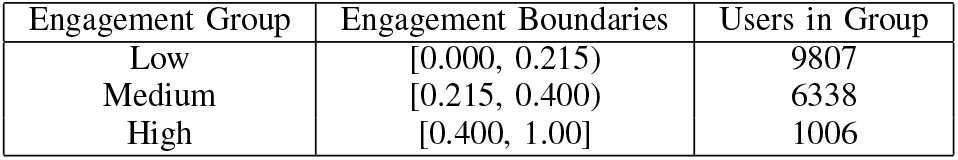
Engagement group boundaries with the number of users in each.

| Engagement Group | Engagement Boundaries | Users in Group |
| --- | --- | --- |
| Low | [0.000, 0.215) | 9807 |
| Medium | [0.215, 0.400) | 6338 |
| High | [0.400, 1.00] | 1006 |

## III. Results

### A. Database Summary

Of the 25,782 users who give approval for their data to be used for research purposes: 3383 report being female, 1997 report being male (the rest are non-reporting). These users have a range of ages, from young children to the elderly, with an average age of 28. The vast majority of users — 59% — that report location come from European nations, with the next highest being: North America —-11% and Asia — 3%. Finally, the median number of seizures reported is only 2 across the whole set of users, with 45% not reporting any at all.

Data on sleep (N=7478) revealed an average sleep quality score of 6.5, while mood data (N=7302) showed an average score of 6.4. The proportion of individuals with sleep scores below the 25th percentile was 22.4%, and those with mood scores below the 25th percentile was 22.7%. Seizure clusters were reported in 19.5% of the sample.

### B. Investigating the Groups

It is clear from the summary of the data that not all variables are being reported, with notable differences between male and female reported patients. Looking at some of the more important variables that may change over time (seizures, mood and sleep scores) between genders and the engagement groups, we find the results in table II. Here we show the percentage of males and females in each group that are reporting seizures, mood and sleep. For each of these engagement groups we would like to understand how engagement changes over time to see if there are any significant differences. For this we look both at an expanding window of time, and fixed windows. For each of these, a time-step of 3 months is used. It should be noted that the data does not have information on when elements of the user’s profile were added. Without knowledge of when features such as birth year, gender, medications etc were first interacted with, we cannot look at PC or MR changes over time in each group. As a result, these metric components are kept constant throughout for each user in a group. Engagement changes, then, entirely come from uploads of sleep and seizure scores which do have dates associated with them.

**TABLE II:** Reporting percentage for key, time-dependent variables between groups and sexes. The largest difference we can see is between the low and medium groups, especially in the case of the non-biologically triggered variables: mood and sleep.

| Group | Sex | Seizures (%) | Mood (%) | Sleep (%) |
| --- | --- | --- | --- | --- |
| Low | Female | 73.48 | 49.91 | 53.78 |
|  | Male | 71.68 | 44.53 | 44.82 |
| Medium | Female | 87.73 | 79.99 | 81.18 |
|  | Male | 87.66 | 74.03 | 74.79 |
| High | Female | 97.71 | 98.67 | 100.00 |
|  | Male | 84.70 | 99.36 | 100.00 |

For an expanding-window, average engagement is calculated for the first 3 months period, and an average found. This is then repeated for the first 6 months, 9 months etc until the maximum number of periods is reached. The results of this are shown in figure 5. Here there is a consistent engagement over time for all groups with a slight downward trend in the higher group and a slight upward trend for the lower engaged group. The standard deviation shows that the variability in engagement is similar across groups but that the medium group tends to drop over time with a spike closer to the later intervals. For the fixed-window, average engagement is calculated for the first 3 month period. 3 months is chosen to reflect common clinical intervals such as: period of time needed before someone is considered seizure free enough to drive [27], how long before seeing whether certain treatments have worked [28], and also to reflect how businesses have typically measured engagement in quarterly (3-month) reviews. However, this method differs by considering each time-step as an independent window, rather than expanding the window by the time-step. This allows us to understand how the engagement progresses over time as isolated windows. The results are shown in figure 6. This shows that in each interval, medium and low stay relatively constant, however highly engaged users experience a significant drop in engagement towards the end of the time spent on the app. There is also much more variability within windows as shown by the standard deviation over time in the high engagement group. This drops to match the values in other groups towards the end.

**Fig. 5:**
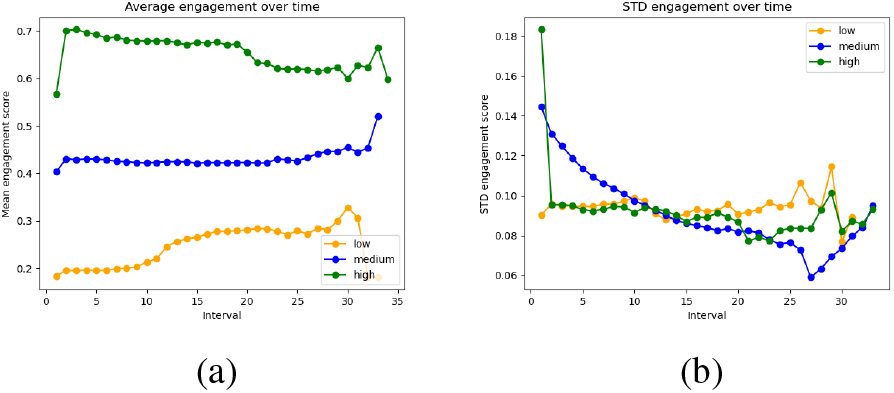
Plots of the (a) average engagement over time as well as the (b) standard deviation of engagement over time for the expanding window. Each “interval” is 3 months.

**Fig. 6:**
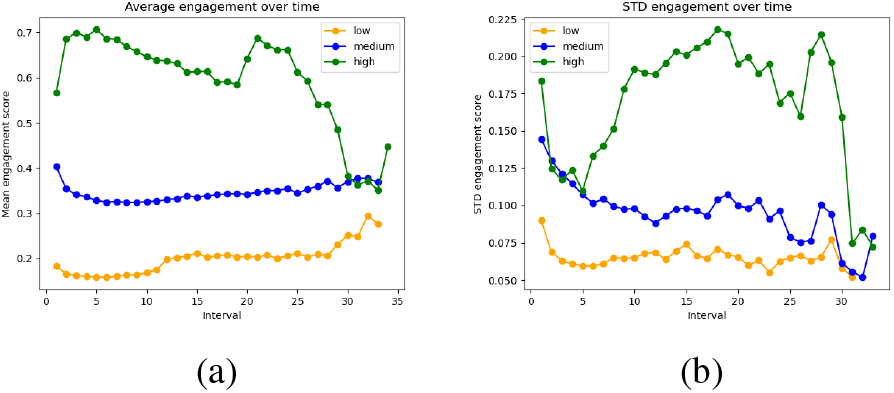
Plots of (a) average engagement over time as well as (b) the standard deviation of engagement over time for the fixed window. Each “interval” is 3 months.

Table III shows the mean, minimum and maximum number of intervals that each group is active on the app. Whilst there is an increase from low to medium, the highly engaged group shows a significantly lower amount of time spent. This could go some way to describe the higher variability in the expanding window, as more people stop using early.

**TABLE III:**
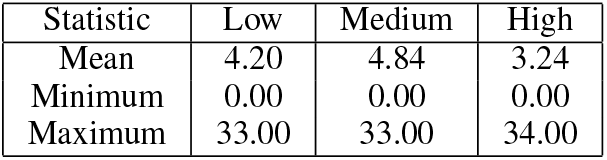
The mean, minimum and maximum intervals the different groups are on the app for. Each interval is 3 months.

We can begin to investigate what makes these groups different by looking at the relative importance of each component of the metric. Figure 7 shows a radar plot of these components with the values for each scaled to how large the component is compared to each group. This shows some interesting features. For almost all components, the highly engaged group is the highest, which is expected. However, the seizure upload regularity component is at 0. The medium engaged group has small values for seizure and sleep upload regularity and the TUR of sleep. profile completeness ratio and medication reminder is high in comparison. For the low engaged group, all values show 0 except the seizure upload regularity which is the highest relative to the other groups. Table IV shows the mean values of each component for each group and table V shows those values after being scaled per feature according to a min-max scaling, defined as: The values in table IV show that regularity is low across all groups. Whilst it may seem that there are vast differences when only considering the radar plot, the largest absolute differences between groups come from medications, how often sleep scores are uploaded (TUR) and how complete the profile is. This is something we see in table II where the largest differences in upload percentages are seen in sleep and mood.

**TABLE IV:** Mean values for each engagement metric component for each group.

| Group | PCR | TUR | $R_{sleep}$ | $R_{seiz}$ | MR |
| --- | --- | --- | --- | --- | --- |
| Low | 0.699 | 0.017 | $2.925 \times 10^{-9}$ | $2.482 \times 10^{-8}$ | 0.006 |
| Medium | 0.831 | 0.162 | $9.689 \times 10^{-9}$ | $1.092 \times 10^{-8}$ | 0.886 |
| High | 0.837 | 0.836 | $1.929 \times 10^{-8}$ | $9.363 \times 10^{-9}$ | 0.830 |

**TABLE V:** Min-max scaled mean values for each engagement metric component for each group.

| Group | PCR | TUR | $R_{sleep}$ | $R_{seiz}$ | MR |
| --- | --- | --- | --- | --- | --- |
| Low | 0.000 | 0.000 | 0.000 | 1.000 | 0.000 |
| Medium | 0.959 | 0.111 | 0.413 | 0.101 | 0.937 |
| High | 1.000 | 1.000 | 1.000 | 0.000 | 1.000 |

**TABLE VI:** Mean doses of the five most popular drugs in the low, medium, and high engagement groups.

| Group | Levetiracetam | Lamotrigine | Lacosamide | Valproic Acid | Carbamazepine |
| --- | --- | --- | --- | --- | --- |
| Low | 1000 | 150 | 100 | 750 | 800 |
| Medium | 1000 | 162.5 | 100 | 750 | 400 |
| High | 1500 | 200 | 300 | 1000 | 1000 |

**Fig. 7:**
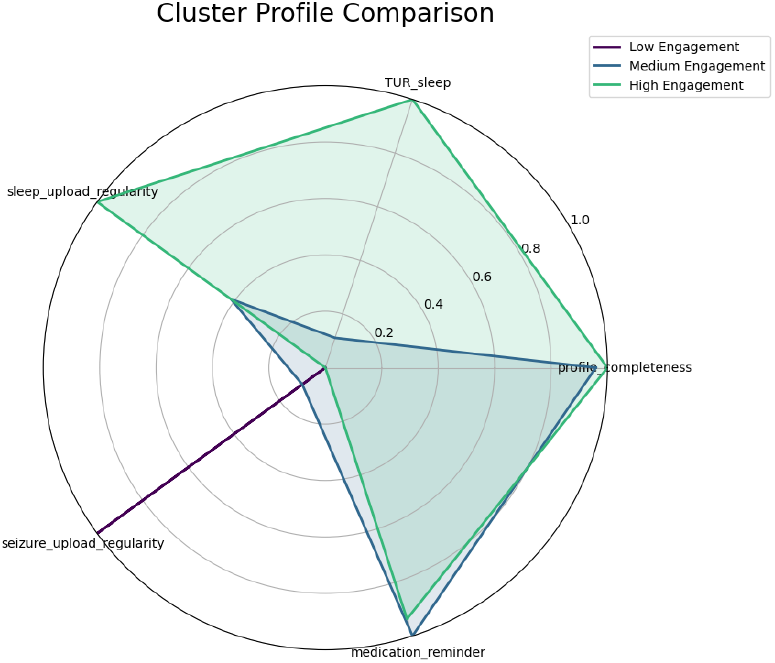
The relative magnitudes of each component of the engagement metric between groups.

Exploratory analyses identified age and medication-related variables as the primary differences between groups. While a range of additional clinical and demographic characteristics were assessed, these did not demonstrate robust associations in the present dataset. This is likely because of missing data and inconsistent uploads. Figure 8a shows birth year vs engagement group. There is a marked difference between the groups, especially the highly engaged group where age skews higher than the others. Median values are indicated by the horizontal lines in the box-and-whisker plots. For the groups, low has a median of 2002, medium 1999, and high 1993. Figure 8b shows a similar trend for the age of diagnosis where the older someone is at the time of learning they have epilepsy, the more likely to be engaged they are. For the groups, low has a median of 11, medium 14, and high 18 years of age. Figure 8c extends this discussion of age to the number of years a patient has had their diagnosis. This shows another positive correlation between group and age, indicating that those that are highly engaged tend to have had more experience with their epilepsy. For the groups, low has a median of 24, medium 27, and high 33 years since diagnosis.

**Fig. 8:**
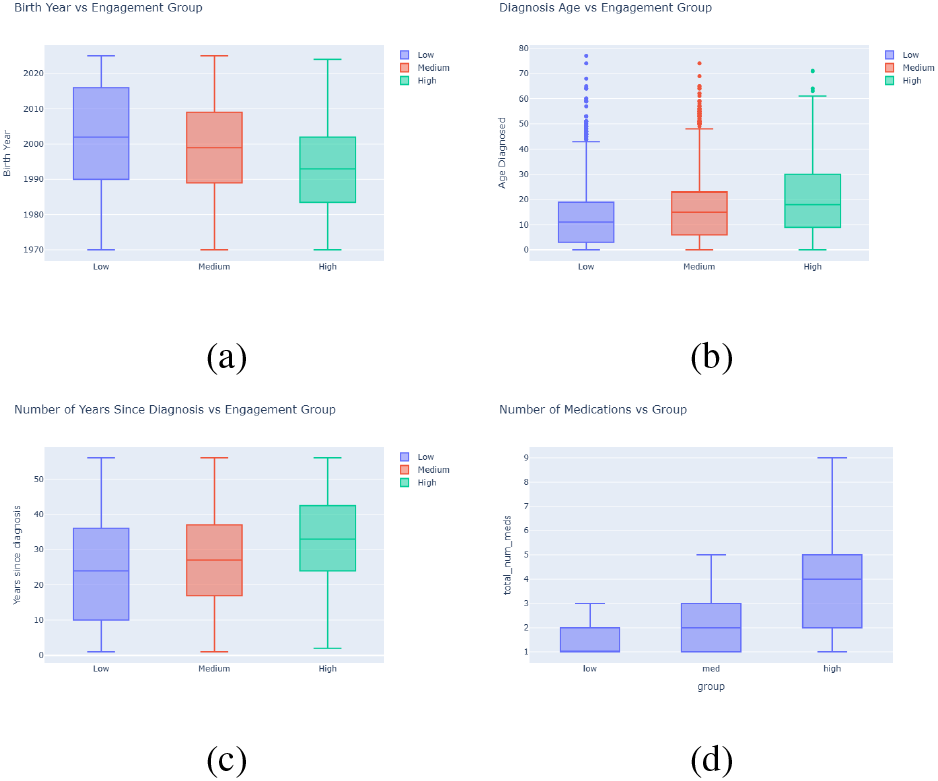
A show of (a) birth year vs engagement group (b) age when diagnosed vs engagement group (c) number of years since diagnosis vs engagement group (d) number of ASMs vs engagement group. Values are taken after 1970.

Finally, medication information is also important for the ongoing treatment of epilepsy. Figure 8d shows the total number of anti-seizure medications (ASMs) attributed to each patient. This includes any ASMs they may have taken in the past. Whilst all patients in the groups report at least 1 ASM, the highly engaged group have a much higher value (4) than medium (2) and low (1). When examining the differences between the ASMs that each group has, there is little difference between the low and medium engaged groups. Both of these groups show the top five most prescribed drugs to be: levetiracetam, lamotrigine, lacosamide, valproic acid and carbamazepine. However, the highly engaged group has more second-line medications, as well as medications that are typically prescribed to users with refractory epilepsy. The top 5 for the highly engaged group are: carbamazepine, brivaracetam, valproic acid, lacosamide and levetiracetam. Table VI shows that for the top drugs shown in medium and low engagement groups, the high engaged group is typically prescribed a higher dose of each.

This shows that not only are the highly engaged group taking more ASMs, they are taking more specialised ASMs and higher doses of the common first-line drugs. These results show clear and clinically relevant differences between the engagement groups.

## IV. Discussion

We showed that an engagement metric based on seizure and sleep score uploads, app profile completeness ratio and whether someone has medication reminders set can split users of a seizure diary application into high, medium and low engagement groups. These clusters are supported by analysing the elbow plot of the data and high silhouette scores.

### A. Engagement Groups

We find that the low engagement group tends to report sleep and mood scores less, though they use the app as a seizure diary somewhat more often than the high and medium engaged groups. Adding to this that all other components of engagement are significantly lower than in the medium or high groups, the low engagement users are likely using this app as a seizure diary only and not fully engaged with other, clinically important elements.

In contrast, the medium engaged group has high profile engagement and medication reminder scores, but lacks in others. Compared to the highly engaged group, we see comparatively low scores for sleep score upload regularity and TUR. With seizure upload regularity also low, this indicates that the medium engaged group are those that may use the app as a seizure diary, but also as a way of remembering medication. They may not, however, have confidence that the app has other clinically useful elements and perhaps are not aware of the importance of things like sleep to epilepsy management (sleep, or the lack thereof, being a common seizure trigger). In support of this, the starkest difference between the percentage reporting of medium and high engaged groups, across male and female responders, is in the percentage reporting mood or sleep scores.

The highly engaged group has similar seizure upload regularity, profile completeness ratio and medication reminder scores as the medium group, but much higher sleep score upload regularity and TUR relative to the low and medium engaged group. This indicates users that are likely more involved in the management and treatment of their epilepsy.

This could be explained by looking at the age and medication-based analysis of the differences between the groups. Starting with the ages, we showed that: the older someone is at time of diagnosis, the older they are and the more years they have known about their epilepsy, the more engaged they tend to be. This may be because being more mature when you get a diagnosis means a user would have more of a hands-on approach to epilepsy management, whereas a younger person receiving a diagnosis may be more likely to be unable to fully understand the seriousness of the condition. Also, younger diagnosis could mean that care is delegated to a guardian, meaning that epilepsy treatment was something done for them, rather than something they are an active part of.

In addition, the higher number of ASMs in the more engaged groups shows that those that have had many drug changes are more likely to be motivated to be engaged in any process that helps their treatment. The fact that doses are higher for common medications, and there are more refractory-type ASMs present in the highly engaged group, shows that the highly engaged users are likely to be those with treatment resistant epilepsy, and also more likely to tolerate possible negative side effects from higher doses of medication in order to achieve a better control of epileptic symptoms. This is a clear sign that more time has been spent with clinicians, therefore more intervention by medical professionals, and so more conversations and information shared in a closed loop between patient and clinician.

As a result of these findings, we can put together a profile of the highly engaged user: someone who is more likely to have had difficulty in treating their epilepsy and is more motivated to try and search for the best resolution they can. In fact, when we look at the engagement over time, whilst we see that the interval-to-interval engagement in all the groups stays reasonably consistent, the number of intervals the highly engaged user spends on average is considerably lower than in the others. We do not see a shift in numbers of users in each engagement bracket over time either, showing that users tend to stay in their engagement groups. This could show that the more engaged a user is, the less they actually need to use the app because they get what is clinically useful and relevant from it and their clinician is able to use the information to prescribe the best treatment.

Consistent with this, Miller et al. [29], analyzing long-term use of the My Epilepsy Diary application, identified four distinct tracking trajectories and demonstrated a marked decline in engagement over time, with the proportion of participants recording at least one diary entry per month decreasing from 86% in the first month to 36% after three years.

It should be also noted that churn rate — the number of users leaving a platform — on applications on iOS and Android can be higher than 50% after a year [30] with health and wellness apps being up to 60% [31]. The latter, however, can drop to 15-25% with clinical backing and habit-forming app elements. This supports a paradigm of teaching those using a health app, such as Helpilepsy, how to effectively use the app, what the outcomes of high engagement with it could be for them, and increasing the use of habit-forming activities such as gamifying elements of the application.

### B. Implications for Digital Epilepsy Monitoring

Previous studies using Helpilepsy® have primarily focused on usability, adherence, retention, and the validity of self-reported data. Zabler et al. [18] reported high usability and acceptability of the platform and emphasized the importance of user training and continued feature development. Similarly, Macea et al. [14] demonstrated moderate long-term adherence and identified reporting burden as a key factor influencing continued use, suggesting that monitoring strategies should be personalized. Dedeken et al. [17] further showed that engagement is associated with demographic and clinical characteristics, including age, sex, and seizure burden, while also demonstrating that integration with healthcare professionals substantially improves retention.

Evidence from other chronic neurological disorders provide additional information. In migraine, electronic diaries have been used to characterize headache trajectories, monitor treatment response, and identify predictors of sustained use, demonstrating that engagement is influenced by both clinical and behavioral factors [32] [33] [34]. Similar findings across chronic diseases highlight the importance of understanding engagement to optimize digital health interventions and improve clinical outcomes. Engagement is a multidimensional concept involving the frequency, consistency, breadth, and persistence of use, while attrition remains a common challenge in real-world settings [35].

The present study applied a multidimensional engagement framework to a large real-world cohort of Helpilepsy® users and found that engagement with electronic seizure diaries is influenced not only by app features but also by patient characteristics and disease complexity. Highly engaged users were generally older, had lived with epilepsy for longer, and reported a greater medication burden, potentially reflecting increased treatment complexity and motivation for self-management.

Future studies should investigate whether engagement can be improved through personalized approaches tailored to clinical characteristics, disease severity, and individual monitoring needs. Given the lower engagement observed among younger users and those with shorter disease duration, targeted on-boarding, educational support, and individualized monitoring strategies may be particularly beneficial. Similarly, reducing user burden through passive data collection, wearable integration, and closer clinician-app interaction represents a promising avenue for future development. Further research is needed to determine whether higher engagement translates into improved clinical outcomes, treatment adherence, and long-term epilepsy management.

### C. Limitations

As mentioned in section III-A, there is a large proportion of people that do not report seizures, mood/sleep scores etc. This reflects the sparsity in the data, with many users only entering a few details or, in some cases, none at all. As a result, it can be difficult to draw concrete conclusions around common avenues of exploration like: auras, seizure triggers, comorbidities etc, for which there is almost no data at all here. We see that seizure and sleep regularity are low for all groups, showing this sparsity: users tend to upload every now and again with no particular pattern. Having a more in-depth onboarding process, emphasising the benefits of uploading more information and more frequently could mitigate these issues and increase the wealth and quality of data available.

The age-based variables found in figure 8 are mathematically collinear, so treating them as separate lines of evidence should be done with the caveat that they may well be correlated. There is not necessarily a direct correlation between them, however, so they are reported here as pseudo-independent pieces of evidence.

It should be noted that, as also mentioned in section III-A, there are demographic differences in the users. There are more than 1.5x more users that report being female than male. This may not reflect the true distribution given the vast majority do not report. In addition, almost 60% of all users come from the EU, reflecting the fact that Helpilepsy is an EU-based application. Pushing the app into other regions is a clear way of decreasing this bias.

### D. Future Work

This analysis has much scope for continuation. First, the sleep scores were chosen as an example of an uploadable feature on the app that is not motivated by seizure occurrence. This could be extended to mood scores, as well as other upload features like: physical health scores (how one’s body rather than mind feels), user-written notes about the day etc. Study into this would determine whether there are other features that could contribute to engagement, as well as whether engagement scores could be generalised to other record modalities e.g. clinic notes. This final point should be investigated by looking into other forms of seizure diaries such as digitising paper diaries kept by patients and seeing whether they too fall into different groups.

Noting the low values for regularity, studying whether removal of this element has an effect on the metric would also be an interesting avenue of exploration. Similarly, varying the weights of the components may yield a more discriminating metric overall. For instance, the TUR of sleep uploads is clearly very different between the groups. This shows that this could be a much more important element than, say, whether someone has a medication reminder set.

Finally, more dense datasets with clinic-aided onboarding for users would yield a richer dataset that could provide more clinically relevant information. Focusing on understanding patient auras, triggers and a more regular uploading of scores would make avenues of investigation such as seizure cycle identification easier to perform. In addition, having more information on the number of clinic dates would help us understand whether this is correlated with higher engagement.

## V. Conclusion

We present here a study into the formation of a novel engagement metric based on data collected from users of the Helpilepsy app from Neuroventis. It is shown that the engagement metric splits the dataset into clear groups of low, medium and highly engaged users, with main differences coming from the TUR of sleep, and age-based investigations of the dataset. It is seen that users are consistently in similar engagement groups over their lifetime use of the app. We go on to show that these groups have significant differences in how old users were when diagnosed, how long they have had epilepsy as well as their age at time of using the app. This indicates a component of maturity when it comes to understanding app engagement. In addition, those in the high engagement groups show more ASMs, as well as more second-line and refractory-type treatments. Also, doses of common drugs such as lamotrigine and carbamazepine are seen to be higher in the most engaged group.

More work is required to increase the validity of these results, with data sparsity being a main issue. However, we are able to put together a profile of the engaged user: one who got an epilepsy diagnosis later in life and who has had many clinic interventions as well as time with the illness. This points to engagement coming as a result of understanding the disease and wanting to be active in being as comfortable as possible whilst experiencing it.

## Data Availability

Data is not available without explicit permission from Neuroventis.

## Notes

All other authors (JD AB) declare no conflicts of interest.

### Competing Interest Statement

L.A & J.S. are employees of Neuroventis, a Cascador Health Company.
MPR received speaker fees from UNEEG medical and consulting fees from UNEEG medical and Lundbeck. MPR received funding support from the NIHR Biomedical Research Centre at the South London and Maudsley NHS Foundation Trust.
P.F.V. has received travel and consultancy fees from UNEEG Medical but declares no nonfinancial competing interests.
All other authors (JD & AB) declare no conflicts of interest.

### Author Declarations

All data was given and approved by the company Neuroventis for use in the study because of data sharing agreements with the Basic and Clinical Neuroscience team at KCL. Because the study involved analysis of existing de-identified data and did not involve recruitment, intervention, or access to identifiable personal information, additional Research Ethics Committee approval was not required under applicable UK research governance regulations.

